# Risk Factors for Intracranial Hemorrhage Transformation and Poor Prognosis after Endovascular Treatment for Acute Large Vessel Occlusion Stroke

**DOI:** 10.1101/2023.05.25.23290563

**Authors:** Lei Ma, Dongfeng Wang, Zhenqiang Li, Gengfan Ye, Pandi Chen, Maosong Chen

## Abstract

**Purpose:** The purpose of this study was to explore the risk factors for hemorrhagic transformation after endovascular treatment for acute large-vessel occlusion stroke and the risk factors for poor prognosis after endovascular treatment.

**Methods:** A total of 132 cases of patients with acute large vessel occlusion stroke who underwent endovascular treatment in the Ningbo Medical Center Lihuili Hospital 1, 2021 to December 31, 2022 were included. According to whether there was intracranial hemorrhage within 72 hours after surgery, the patients were divided into hemorrhage transformation(HT) group and non-hemorrhage(no HT) group. The two groups were compared in terms of gender, age, history of hypertension, history of atrial fibrillation, and other factors to determine whether there were statistical differences.

**Results:** Among the 132 included patients, 30 cases had hemorrhage transformation and 102 cases were in the non-hemorrhage group. Multivariate logistic regression analysis showed that the ASPECTS score on admission and the NLR within 24 hours after surgery were independent risk factors for hemorrhage transformation. The rate of good prognosis (MRS score 0-2) at discharge was 31.2%, and multivariate logistic regression analysis showed that advanced age, history of hypertension, and NIHSS score were independent risk factors for poor prognosis.

**Conclusion:** The NLR within 24 hours after endovascular treatment is an independent risk factor for hemorrhage transformation in acute large vessel occlusion stroke and can be used as one of the predictive indicators. Advanced age, history of hypertension, and NIHSS score are independent risk factors for poor prognosis after endovascular treatment for acute large vessel occlusion stroke.

## 1. Background

With the rapid development of China’s economy and the continuous improvement of living standards and medical conditions, the aging trend of the population is becoming more and more obvious, and the incidence of acute ischemic stroke is on the rise. Ischemic stroke is mainly caused by intracranial vascular occlusion leading to blood circulation disorders, ischemia, and hypoxia, which can result in local brain tissue softening and necrosis. It mostly occurs in middle-aged and elderly people and is a common cerebrovascular disease. Due to its high disability, mortality, and recurrence rates, ischemic stroke is an important disease that threatens human health^1^. Endovascular treatment is an effective method for treating large vessel occlusion in acute ischemic stroke. With the development of endovascular treatment technology, although the revascularization rate of patients has been significantly improved, the prognosis of the disease has not been significantly improved, and even some studies have shown that the incidence of hemorrhagic transformation in patients after endovascular treatment has increased significantly^2^. Hemorrhagic transformation is a known complication after reperfusion, and the substantial hematoma with mass effect has a high incidence and mortality rate. In recent years, studies have shown that 9%-49.5% of acute ischemic stroke patients who undergo endovascular treatment experience hemorrhagic transformation, and 2%-16% of these patients have symptomatic cerebral hemorrhage^3-8^. Predictive factors for hemorrhagic transformation in acute ischemic stroke patients who receive intravenous thrombolysis have been widely studied, but there is still little literature on hemorrhagic transformation in endovascular treatment^9^. This study collected clinical data of patients with acute large vessel occlusion ischemic stroke who underwent endovascular treatment in the neurosurgery, neurology, and NICU departments of two campuses of our hospital from January 1, 2021, to December 31, 2022, and analyzed the risk factors for hemorrhage transformation after endovascular treatment. We hope that this study will provide help for the diagnosis and treatment of patients with acute large vessel occlusion ischemic stroke. This study has been approved by the ethics committee (approval number: KY2023SL138-01) and has received funding from the Ningbo Municipal Key Medical Discipline.

## 2. Data and Methods

### 2.1 Study Objectives

This study collected clinical data of 138 patients with acute large vessel occlusion ischemic stroke who underwent endovascular treatment at the Li Huili Hospital East Campus and Xingning Campus of Ningbo Medical Center from January 1, 2021, to December 31, 2022. Six cases were excluded due to incomplete clinical data, and 132 cases were included in the analysis, including 89 males and 43 females. The age range was 36-92 years (mean age 67.3 ± 12.9 years). Inclusion criteria were: (1) age ≥18 years; (2) clear diagnosis of acute ischemic stroke; (3) confirmation of intracranial large artery occlusion by computed tomographic angiography (CTA), magnetic resonance angiography (MRA), or digital subtraction angiography (DSA); (4) National Institute of Health stroke scale (NIHSS) score ≥6 points; (5) onset time ≤6h, or 6-16h: meeting the DAWN/DEFUSE 3 criteria: low perfusion volume/infarct core>1.8 and maximum infarct core<70ml, or 16-24h: meeting the DAWN criteria: (1) onset 6-24 hours, (2) CT angiography confirmed internal carotid artery intracranial segment and middle cerebral artery M1 segment occlusion, (3) age≥80 years, NIHSS≥10, infarct volume <21ml, or age <80 years, NIHSS≥10, infarct volume <31ml, or age <80 years, NIHSS ≥20, 31ml<infarct volume<51ml; (6) Alberta Stroke Program Early CT Score (ASPECTS) ≥6 points; (7) other patients who did not meet the above criteria but could still benefit from endovascular treatment after comprehensive evaluation. Exclusion criteria were: (1) patients with incomplete clinical and imaging data during the perioperative period; (2) preoperative Modified Rankin Scale (mRs) score ≥3 points. All patients signed an informed consent form for the surgery.

### 2.2 Treatment methods

If the patient meets the indications for intravenous thrombolysis and there are no contraindications, alteplase is administered for thrombolysis treatment prior to surgery. Endovascular treatment includes catheter aspiration, stent retrieval, balloon angioplasty, and stent implantation. After successful local or general anesthesia (local anesthesia before intra-arterial therapy with remifentanil 20ml/h sedation; general anesthesia after intra-arterial therapy), an 8F guiding catheter is inserted into the proximal part of the lesion site in the internal carotid artery, middle cerebral artery, or vertebral artery through a 0.035-inch guide wire. The guide wire is removed, and a 0.014-inch micro-guide wire and microcatheter are inserted along the guiding catheter to the vascular lesion site, and carefully passed through the lesion site to the distal normal vessel. The micro-guide wire is removed, and an angiography is performed from the microcatheter to confirm that it is located in the vascular lumen and the distal blood vessel is unobstructed. The retrieval stent is sent to the vascular lesion site through the microcatheter and released, and the retrieval stent and thrombus are intertwined. The middle catheter is delivered to the proximal end of the retrieval stent and ensures that the middle catheter passes important vascular branches. The retrieval stent is withdrawn while the middle catheter is aspirated with a 30 ml empty syringe, and the stent and thrombus are removed. In addition to using a retrieval stent in combination with a middle catheter for retrieval, a thrombus can also be directly aspirated with a suction catheter depending on the vascular condition. Successful vascular recanalization is defined as achieving a modified Thrombolysis in Cerebral Infarction (mTICI) grade of 2b/3. If the first retrieval fails, the same retrieval steps can be repeated up to five times. If the post-retrieval angiography reveals severe stenosis of the vascular lesion and the mTICI grade cannot reach 2b/3, balloon angioplasty alone or in combination with intracranial arterial stent implantation may be performed, and antiplatelet therapy with clopidogrel is maintained during the surgery.

### 2.3 Imaging test

Imaging examinations include head CT, head CTA, head CTP, DSA, etc. The imaging data of all patients were independently reviewed by a senior neurosurgery specialist and a senior radiology specialist. In case of disagreement, a third senior neurosurgery specialist reviewed the images and the differences were resolved through discussion to reach a consensus.

### 2.4 Data collected

The collected clinical data include: patient age, sex, onset time, underlying diseases (such as hypertension, diabetes, hyperlipidemia, atrial fibrillation, coronary heart disease, history of stroke, etc.), NIHSS score at admission, ASPECTS score, blood pressure, history of anticoagulant treatment, history of antiplatelet treatment, DNT (time from admission to start of intravenous thrombolysis), DPT (time from admission to successful vascular puncture), DRT (time from admission to blood vessel recanalization), blood glucose level at admission, blood routine within 24 hours before and after surgery (including NLR (neutrophil to lymphocyte ratio), hemoglobin level, lymphocyte to platelet ratio), triglycerides, LDL-C, albumin, operation time, intracranial atherosclerotic stenosis, method of thrombus retrieval, postoperative mTICI grading, number of thrombus retrieval attempts, head CT scan within 72 hours after surgery to determine the presence of hemorrhagic transformation. If the presence of intracranial hemorrhage cannot be accurately judged due to interference from contrast agent extravasation, head CT scans may be repeated at 24-hour intervals to assist in the diagnosis. MRS score at discharge and 30 days after discharge. Patients were divided into hemorrhagic and non-hemorrhagic groups based on CT scan results.

### 2.5 Statistical analysis

The collected data were analyzed using SPSS 26.0 statistical software. Continuous variables were expressed as means ± standard deviation 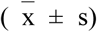, and the t-test was used for inter-group comparisons. Categorical variables were presented as numbers or percentages. Variables with P < 0.1 in univariate analysis were included in the multivariate logistic regression model to evaluate their independent predictive ability for hemorrhagic transformation after EVT. The data from the logistic regression analysis were presented as odds ratios (OR) and their corresponding 95% confidence intervals. The predictive value was assessed using receiver operating characteristic (ROC) curve analysis. A P value of < 0.05 was considered statistically significant.

## 3. Results

### 3.1 Logistic regression analysis of hemorrhagic transformation after endovascular treatment for acute large vessel occlusion stroke

A comparison between the bleeding and non-bleeding groups revealed statistically significant differences in NLR (OR=1.105, 95% CI=1.036-1.179, P=0.003) and ASPECTS scores (OR=0.784, 95% CI=0.640-0.962, P=0.0194) within 24 hours after surgery, with P<0.05. Detailed baseline and clinical characteristics are organized in Table 1.

### 3.2 Logistic regression analysis for poor prognosis (MRS score > 2) at discharge

The comparison between the bleeding and non-bleeding groups showed that there were statistically significant differences (P<0.05) in age (OR=1.069, 95%CI=1.011-1.131, P=0.020), history of hypertension (OR=0.335, 95%CI=0.114-0.981, P=0.046), and NIHSS score (OR=1.106, 95%CI=1.010-1.211, P=0.029). Detailed baseline and clinical characteristics are organized in Table 2.

## 4. Discussion

Clinical studies have shown that endovascular treatment has a definite advantage in the treatment of acute ischemic stroke with large-vessel occlusion, but although the patient’s blood vessel recanalization rate has been significantly improved, studies have shown that the incidence of hemorrhagic transformation in patients after endovascular treatment has increased significantly^2^. ZHANG et al. found that the incidence of symptomatic intracranial hemorrhage in the real world was 13.8%^10^. In this study, the rate of hemorrhagic transformation after endovascular treatment was 22.7%, which is similar to previous studies. Logistic regression analysis of the factors contributing to hemorrhagic transformation after endovascular treatment revealed that the ASPECTS score (OR=0.784, 95%CI=0.640-0.962, P=0.0194) was significantly associated with hemorrhagic transformation, which is consistent with previous literature reports ^11, 12^. The neutrophil-to-lymphocyte ratio (NLR) within 24 hours after surgery (OR=1.105, 95%CI=1.036-1.179, P=0.003) was also an independent risk factor for hemorrhagic transformation, with a statistically significant difference (P<0.05). ROC curve analysis revealed an AUC of 79.1% (P<0.01, 95%CI=0.705-0.877), and the optimal cutoff value was 0.551, with a sensitivity of 76.7% and specificity of 78.4%. Therefore, the NLR level within 24 hours after endovascular treatment can be used as a sensitive predictor of hemorrhagic transformation. Normal brain tissue is devoid of white blood cells. However, during cerebral ischemia and hypoxia, white blood cell infiltration can be detected. Studies have demonstrated a significant increase in white blood cell adhesion to small cerebral veins within one hour of cerebral ischemia-reperfusion^13^. NLR can better reflect different inflammatory states of the body, and is one of the recognized indicators of systemic inflammatory response due to its advantages of easy acquisition, rapid detection, high sensitivity, low cost, and non-invasiveness^14^. NLR changes with the progression of certain diseases, and this change is consistent with the severity of certain diseases. Its elevated level is accompanied by an increase in neutrophils and a decrease in lymphocytes, indicating an unbalanced interaction between central and peripheral inflammation associated with stroke ^15^. Further research is needed to elucidate the specific mechanism, but NLR has the advantages of low detection cost, low technical requirements, easy acquisition, and suitability for use in primary care hospitals. Compared with previous studies ^11, 12, 16, 17^, this study did not find that atrial fibrillation, blood glucose levels, age, hyperlipidemia, etc. were risk factors for hemorrhagic transformation, which may be related to insufficient sample size. Multivariate logistic regression analysis showed that age (OR=1.069, 95%CI=1.011-1.131, P=0.020), history of hypertension (OR=0.335, 95%CI=0.114-0.981, P=0.046), and NIHSS score (OR=1.106, 95%CI=1.010-1.211, P=0.029) were independent risk factors for poor prognosis (MRS>2) at discharge. These factors can be used as reference indicators for possible poor prognosis after endovascular treatment for acute large-vessel occlusion stroke. The rate of good prognosis (MRS score of 0-2) at discharge in this study was 31.2%. After active rehabilitation and 30-day follow-up by phone, the rate of good prognosis increased to 41.7%. This indicates that active rehabilitation treatment is helpful in improving prognosis. The proportion of patients without hemorrhage who had an MRS score of ≤2 at discharge was 38.2%, which was better than the hemorrhage group’s 10%. This may be the reason why the MRS score at discharge was worse than in other studies^18^.

## 5. Limitations

This study has limitations due to the influence of confounding factors such as sample size, selection bias, and treatment methods, which have some limitations. Therefore, the individual predictive effect of postoperative hemorrhagic transformation after endovascular treatment of acute large vessel occlusion stroke is still in the early exploration stage and requires further verification with larger sample size data. In the future, with the increase in sample size, the study will focus on the impact of clinical indicators on hemorrhagic transformation.

## 6. Conclusion

Within 24 hours after surgery, the neutrophil-to-lymphocyte ratio (NLR) is an independent risk factor for hemorrhagic transformation after endovascular treatment for acute large-vessel occlusion stroke, with an AUC of 79.1%, P<0.01, 95%CI=0.705-0.877, and the optimal cut-off value of 0.551. At this cut-off value, the sensitivity was 76.7% and the specificity was 78.4%. NLR can be used as one of the predictive indicators for hemorrhagic transformation after endovascular treatment for acute large-vessel occlusion stroke. It has the advantages of low detection cost, low technical requirements, and easy acquisition, making it suitable for promotion and application in primary hospitals. Age, history of hypertension, and NIHSS score are independent risk factors for poor prognosis after endovascular treatment for acute large-vessel occlusion stroke and can be used as auxiliary indicators for predicting poor prognosis.

## Data Availability

All data produced in the present study are available upon reasonable request to the authors

## 7. Acknowledgments

## ROC curve for NLR within 24 hours after EVT in cases with hemorrhagic transformation(Figures1)

**Figures1.**
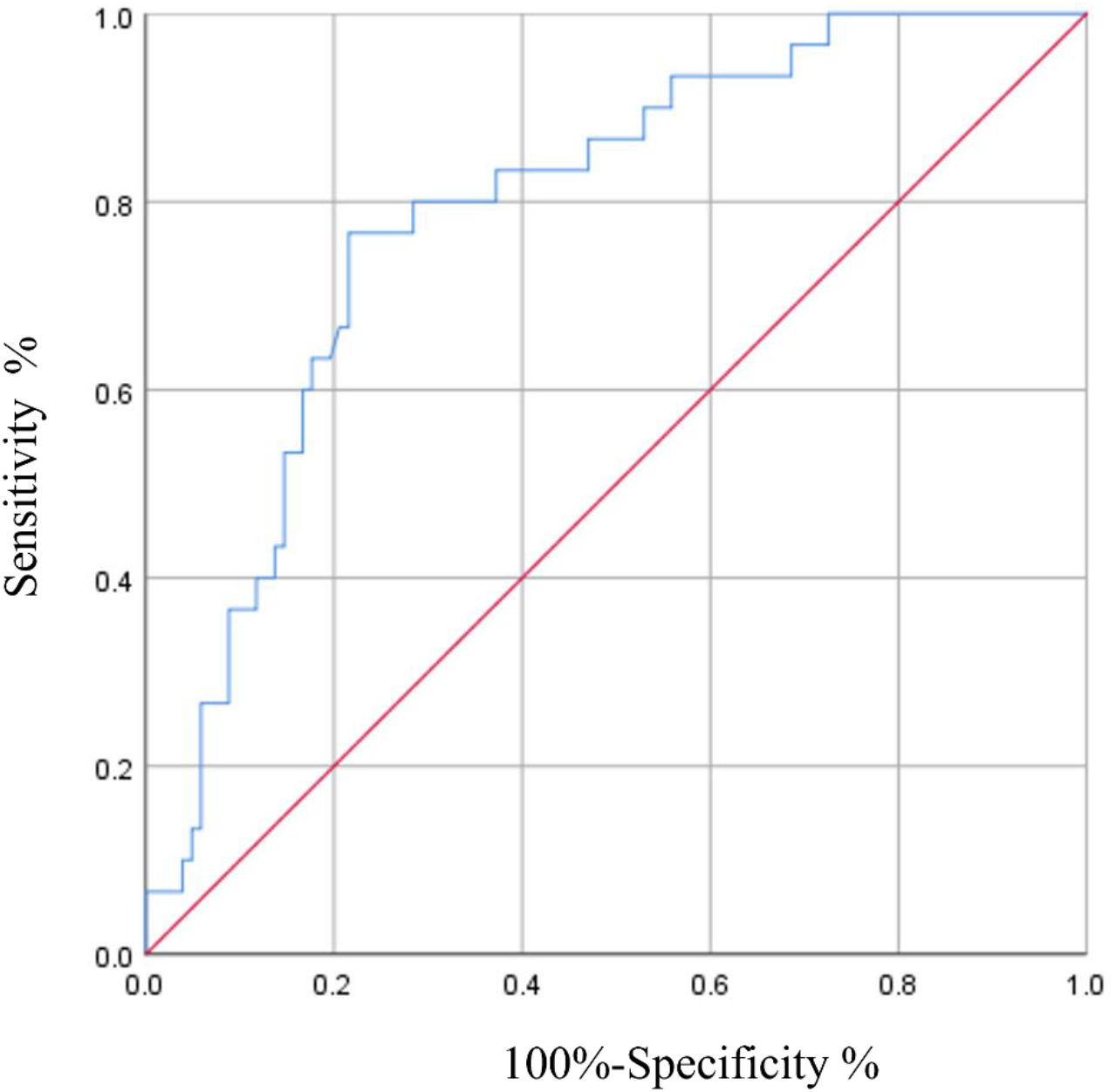
AUC=79.1%,P<0.01,95%CI=0.705-0.877,the optimal cutoff value was 0.551, with a sensitivity of 76.7% and a specificity of 78.4%.

